# County Level Contributors to Geographic Variation in Medicare FFS Stroke Hospitalization Rates: A Cross-Sectional Study

**DOI:** 10.1101/2025.11.18.25340537

**Authors:** Raed Hailat, Mohamed Ridha, Matthew Gusler, Chun Chieh Lin, Kevin Kerber, Jeffrey Wing, James F. Burke

**Affiliations:** The Ohio State University Wexner Medical Center, Department of Neurology; The Ohio State University, Department of Epidemiology

## Abstract

**Background:** Granular regional stroke incidence data in the US is lacking. We sought to identify factors associated with county-level hospitalization rates and counties with hospitalization rates above or below expectation using publicly available data.

**Methods:** This cross-sectional study is based on the analysis of county-level three-year average stroke hospitalization rates (principal ICD-10 I63, I65-I66) per 100,000 population among Medicare fee-for-service (FFS) beneficiaries from 2018-2020 using data from the CDC’s Interactive Atlas of Heart Disease and Stroke (ATLAS) and other sources. ATLAS provided reliable data on 3,198 (98.6%) counties and county-equivalents. Linear mixed models were fitted to investigate six sets of factors (Total of 61) associated with hospitalization rates in a serial additive stepwise fashion (i.e., demographics, overall population vascular risk factors, risk factor treatment, health delivery and access, environmental features, and socioeconomic status). We reported on the predicted hospitalization rates, marginal R^2^ of the fixed effects, the most impactful factors using average marginal effects, and characterized proportional difference between crude and predicted hospitalization rates.

**Results:** The cohort of 3,198 counties and county-equivalents had a mean stroke hospitalization rate of 11.2 per 100,000 (SD= 2.6). Mean characteristics of included counties: 19.4% age ≥65 years, 73% white, 7.6% coronary heart disease (CHD) prevalence, 38% hyperlipidemia prevalence, and 5.7 primary care physicians per 10,000. In the fully adjusted model, between-county unexplained variation remained moderately high (R^2^= 0.57). The most impactful factors associated with stroke hospitalization rates were prevalence of CHD, hypertension, smoking, nonadherence to antihypertensive medication, and the elevation of the county above sea level. Counties in the northwest United States generally had lower than expected hospitalization rates.

**Conclusions:** Considerable unexplained county-level variance in stroke hospitalization rates exists after accounting for a wide variety of known and potential predictors. Future work to clarify the mechanism of known predictors and explain variance may inform stroke mechanisms and interventions to improve systems of care.

## Introduction

In the US, stroke incidence is estimated at 800,000 patients yearly.^1^ This type of global estimation is potentially insensitive to important regional differences in incidence which may be clinically and societally important.^2^ The lack of regional granularity limits our capacity to explore ecological stroke risk factors and to use epidemiologic data to tailor stroke programs and policies towards the specific needs in a community.^3–6^ One strategy to assess regional variation in stroke incidence is to use readily available stroke hospitalization rates, which reflect two underlying factors: stroke incidence and the propensity to seek acute medical care for stroke symptoms.^7, 8^

Regional variation in stroke hospitalization rates is known to exist with evidence suggesting that differences in population characteristics, health risk profile, socioeconomic status, and access to care contribute to this variation.^4, 6, 9^ For example, a recent report by the Centers for Disease Control and Prevention (CDC) demonstrated that counties in the top quartile of stroke hospitalization rates for Black older adults clustered in the Midwest, Northeast, and South, in contrast to stroke hospitalization rates in White older adults that concentrated in the Stroke Belt, specifically the Mississippi Delta region and into Oklahoma and Texas.^4^ In addition, compared to regions with low hospitalization rates, regions with high stroke hospitalization rates were associated with lower median household income, high school education, and number of hospitals at the county level.^9^ Thus far, no study has provided a comprehensive assessment of this variation using an expanded set of factors or outside a binomial stroke hospitalization rate clustered framework.

To address this gap, we designed a hypothesis generating study using county-level Medicare fee-for-service data and publicly available data from the CDC and other sources to characterize regional variation in stroke hospitalization rates, explore predictors of hospitalization rates, and examine the gap between measured and predicted rates.

## Methods

### Goal and hypothesis

Our primary hypothesis was that ecological factors would explain only a small proportion of the heterogeneity of county-level stroke hospitalization rates. Our goal was to explore a wide variety of potential stroke-related factors to generate hypotheses on modifiable stroke risk factors and simultaneously explore the influence of ecological and systems factors on hospitalization rates.

### Study population and Data

This cross-sectional study is based on the analysis of county-level three-year average ischemic and hemorrhagic stroke hospitalization rates (principal ICD-10 I63, I65-I66) per 100,000 population among Medicare FFS beneficiaries from 2018-2020 using publicly available data from the CDC Interactive Atlas of Heart Disease and Stroke (ATLAS).^10^ The ATLAS is an online data resource that both collates national data on relevant constructs (i.e., stroke, demographics, stroke risk factors, social determinants of health, proximity to care) and allows users to easily create and customize local-level maps.^10^ The ATLAS combines data from multiple data sources to this end, including: (1) Medicare FFS part A and D administrative files (stroke hospitalizations), (2) the CDC’s Behavioral Risk Factor Surveillance System (BRFFS)-a nationally representative telephone survey that collects state data about U.S. residents regarding their health-related risk behaviors, chronic health conditions, and use of preventive services, (3) Health Resources and Services Administration Area Health Resources Files (AHRF)-a national data source for information on health facilities, health professions, health status, health training programs, geographic measures, economic indicators, and socioeconomic and environmental attributes, (4) US-Census Bureau, (5) American Hospital Association, (6) American Medical Association, and (7) stroke certification agencies (i.e., DNV-GL, Healthcare Facilities Accreditation Program (HFAP), and The Joint Commission).^10^

### Stroke hospitalization rates

ATLAS hospitalization rates are age-standardized (using the 2000 US standard population) and spatially smoothed using a local empirical Bayes algorithm.^11^ Hospitalization rates are calculated using Centers for Medicare and Medicaid Services Medicare Provider Analysis and Review (MEDPAR) file, Part A.^10^ During the study period the database provided data on 98.6% (n=3,198) of counties and county-equivalents in the US.^10^

### County-level stroke predictors

The county-level data obtained directly from ATLAS and its data providers included the following six sets of predictors that may be associated with stroke hospitalization rates including: (1) demographic composition, (2) stroke risk factors/population health panel, (3) treatment of medical risk factors, (4) healthcare delivery and access, (5) environmental features, and (6) socioeconomic status. Detailed information on the included predictors is found in Table S1 in the supplement. In addition to the ATLAS provided data we included data from the National Cancer Institute-Surveillance, Epidemiology, and End Results Program (SEER), Centers for Medicare and Medicaid Services (CMS), HDPulse platform-an ecosystem of minority health and health disparities resources from National Institute on Minority Health and Health Disparities (NIMHD), The U.S. Bureau of Economic Analysis, and Area Health Resources Files-Health Resources and Services Administration data warehouse.

This study used publicly available county level data and thus did not fall under the guidelines for human subjects research by The Ohio State University Institutional Review Board. This study followed Strengthening the Reporting of Observational Studies in Epidemiology (STROBE) reporting guidelines.

### Statistical Analysis

Our statistical analysis aimed to characterize regional variation in county-level stroke hospitalization rates, identify factors that influence county-level hospitalization rates, and characterize residual regional variation in county-level stroke rates after accounting for probable predictive factors. The unit of analysis was county-level.

Due to the low rate of missingness in potential predictors (0.3%-4.5%, see Table S2 in the supplement), multiple imputation of 10 iterations was used to generate missing values and generate pooled analytical estimates. Multiple imputation used fully conditional specification methodology and included all the predictors used in the analysis. We reported on descriptive statistics (i.e., proportions or mean and medians, SD, linear mixed model estimates and their confidence intervals, association and p-value) pre- and post-multiple imputation of the included predictors using bivariate analysis. Pre-imputation (referred to as complete case analysis) descriptive statistics are reported on in Table S2.

Linear mixed models were fitted to investigate the six sets of factors associated with county-level hospitalization rates in a serial additive stepwise fashion (Table S3). We opted to use this to explore the additive impact of each set of predictors on the predicted hospitalization rates and the unexplained heterogeneity before we report on individual predictors. We started by fitting an empty model predicting stroke hospitalization. In every serial additive step, we calculated the marginal R^2^ of the fixed effects and reported the predicted hospitalization rates descriptive statistics using box plots. Multivariate model estimates of stroke hospitalizations with sequentially added risk factors are reported on in the supplement (Table S4). To characterize the quantitative positive or negative potential effect of the most impactful factors on hospitalization rates, we used the 10^th^ and 90^th^ percentile value of the significantly associated predictors average marginal effects range (90^th^ percentile*predictor model estimate – 10^th^ percentile*predictor model estimate) in the fully adjusted model. Finally, we characterized county level hospitalization rates proportional variation between the predicted and observed values ((observed – predicted)/observed*100) visually using a map for the fully adjusted model. The generated map will characterize the residual variation that isn’t accounted for by the model in a heat map design that designates counties to have above or below expected hospitalization rates. All analyses were conducted in SAS, software v9.4 (Cary, NC). Mapping was done using ArcGIS Pro v3.5.1. Two-sided testing at *P* < .05 indicated significance.

## Results

### Characteristics of Counties and County Equivalents

In a cohort of 3,198 counties and county equivalents representing 98.6% of the US in 2018-2020, the population had a mean prevalence of 19.4% above age 65 years with 65–69-year-olds constituting 6.3% (SD 1.4%), 73.1% (SD 22.8%) white, 7.6% (SD 1.5%) with coronary heart disease (CHD), 38.1% (SD 3.3%) with hyperlipidemia, 8.6% (SD 1.6%) with diabetes, 37.7% (SD 5.5%) reported taking antihypertensive medications, 27.9% (SD 6.6%) were obese, and 18.0% (SD 4.1%) current smoke tobacco (Table 1). Of the included counties, 62.4% were rural, 87.4% included medically underserved areas, and 39.2% did not have a specific primary economic sector (Table 1).

**Table 1:** County level (N= 3,198) descriptive univariate statistics of the risk factors and their association with stroke hospitalization rates using the multiple imputed data.^

| Variable^^ | Description | Mean (SD) or<br>N (%) | Percentile |  | Estimate | 95% CI | p-value |
| --- | --- | --- | --- | --- | --- | --- | --- |
|  |  |  | 10<sup>th</sup> | 90<sup>th</sup> |  |  |  |
| Demographics |  |  |  |  |  |  |  |
| Sex | Percent male of<br>population | 50.0 (2.8) | 47.6 | 52.8 | -0.067 | -0.100 – -0.034 | <0.01 |
| Age 65-69 | Percent of population | 6.3 (1.4) | 4.7 | 8.1 | -0.410 | -0.473 – -0.347 | <0.01 |
| Age 70-74 | Percent of population | 5.1 (1.3) | 3.6 | 6.7 | -0.270 | -0.339 – -0.201 | <0.01 |
| Age 75-79 | Percent of population | 3.5 (1.0) | 2.4 | 4.7 | -0.398 | -0.490 – -0.307 | <0.01 |
| Age 80-84 | Percent of population | 2.3 (0.7) | 1.5 | 3.1 | -0.802 | -0.933 – -0.672 | <0.01 |
| Age 85-89 | Percent of population | 1.3 (0.5) | 0.8 | 1.9 | -1.565 | -1.760 – -1.371 | <0.01 |
| Age ≥89 | Percent of population | 0.9 (0.4) | 0.5 | 1.31 | -2.386 | -2.620 – -2.152 | <0.01 |
| Race White (non-Hispanic) | Percent of population | 73.1 (22.8) | 40.0 | 93.9 | -0.006 | -0.010 – -0.002 | <0.01 |
| Race Black (non-Hispanic) | Percent of population | 8.5 (14.1) | 0.1 | 28.7 | 0.078 | 0.072 – 0.084 | <0.01 |
| Race Asian (non-Hispanic) | Percent of population | 1.3 (2.7) | 0.0 | 3.0 | -0.069 | -0.103 – -0.035 | <0.01 |
| Race American<br>Indians or<br>Alaskan Native<br>(non-Hispanic) | Percent of population | 1.5 (7.0) | 0.0 | 2.0 | -0.002 | -0.016 – 0.011 | 0.72 |
| Race Native<br>Hawaiian or | Percent of population | 0.1 (0.4) | 0.0 | 0.1 | -0.402 | -0.631 – -0.172 | <0.01 |
| Pacific Islander (non-Hispanic) |  |  |  |  |  |  |  |
| Race more than one (non-Hispanic) | Percent of population | 2.9 (1.9) | 1.2 | 4.8 | 0.096 | 0.048 – 0.144 | <0.01 |
| Race other (non-Hispanic) | Percent of population | 0.5 (0.4) | 0.2 | 0.9 | -0.228 | -0.465 – 0.009 | 0.06 |
| Race Hispanic | Percent of population | 12.0 (19.4) | 1.6 | 30.0 | -0.033 | -0.037 – -0.028 | <0.01 |
| Total population | Number of inhabitants in ten thousand | 9.4 (28.8) | 0.5 | 19.7 | 0.0002 | -0.003 – 0.003 | 0.92 |
| Rurality (non-metro counties) | Yes | 1,996 (62.4%) | No | Yes | -0.732 | -0.920 – -0.545 | <0.01 |
|  | No | 1,202 (37.6%) |  |  | ref |  |  |
| Stroke risk factors/population health panel |  |  |  |  |  |  |  |
| Coronary heart disease | Percent of population | 7.6 (1.5) | 5.6 | 9.5 | 0.306 | 0.246 – 0.367 | <0.01 |
| High cholesterol | Percent of population | 38.1(3.3) | 33.7 | 42 | 0.151 | 0.123 – 0.178 | <0.01 |
| Hypertension (antihypertensive medication use) | Percent of population | 37.7 (5.5) | 31 | 44.8 | 0.210 | 0.195 – 0.225 | <0.01 |
| Diabetes | Percent of population | 8.6 (1.6) | 7 | 10.7 | 0.656 | 0.601 – 0.711 | <0.01 |
| Obesity | Percent of population | 27.9 (6.6) | 18.4 | 36.4 | 0.062 | 0.048 – 0.076 | <0.01 |
| Physical inactivity | Percent of population | 19.3 (4.0) | 14.3 | 24.8 | 0.207 | 0.186 – 0.229 | <0.01 |
| Smoking | Percent of population | 18.0 (4.1) | 13.2 | 23.2 | 0.324 | 0.305 – 0.344 | <0.01 |
| Treatment of medical risk factors |  |  |  |  |  |  |  |
| Total Medicare reimbursement per enrollee | Mean of reimbursements in thousands among CVD patients | 22.3 (4.0) | 18.6 | 26.7 | 0.129 | 0.107 – 0.152 | <0.01 |
| Self-reported medication nonadherence to blood pressure medication | Percent of hypertensive population (Medicare part D) | 21.7 (4.2) | 16.4 | 27.1 | 0.207 | 0.186 – 0.227 | <0.01 |
| Self-reported medication nonadherence to cholesterol lowering medication | Percent of hypercholesterolemia population (Medicare part D) | 15.3 (3.6) | 11.6 | 19.2 | 0.150 | 0.125 – 0.175 | <0.01 |
| Healthcare delivery and access |  |  |  |  |  |  |  |
| Hospital | Per 100,000 population | 5.5 (9.5) | 0.0 | 13.3 | -0.075 | -0.085 – -0.066 | <0.01 |
| Hospitals with neurological services | Per 100,000 population | 0.6 (1.7) | 0.0 | 1.7 | -0.039 | -0.095 – 0.016 | 0.17 |
| Hospitals with CT | Per 100,000 population | 3.6 (7.7) | 0.0 | 9.4 | -0.072 | -0.083 – -0.060 | <0.01 |
| Hospitals with medical school affiliation | Per 100,000 population | 0.5 (2.0) | 0.0 | 1.4 | -0.084 | -0.129 – -0.039 | <0.01 |
| Hospitals with tele stroke care | Per 100,000 population | 0.9 (3.1) | 0.0 | 2.5 | -0.037 | -0.066 – -0.007 | 0.01 |
| Certified stroke centers | Per 100,000 population | 0.3 (0.7) | 0.0 | 1.0 | 0.351 | 0.228 – 0.473 | <0.01 |
| Acute stroke ready hospitals | Per 100,000 population | 0.04 (0.37) | 0.0 | 0.0 | 0.207 | -0.042 – 0.455 | 0.10 |
| Primary stroke | Per 100,000 | 0.2 (0.6) | 0.0 | 0.8 | 0.402 | 0.260 – 0.545 | <0.01 |
| centers | population |  |  |  |  |  |  |
| Thrombectomy capable stroke centers | Per 100,000 population | 0.001 (0.03) | 0.0 | 0.0 | 2.096 | -1.083 – 5.275 | 0.20 |
| Comprehensive stroke centers | Per 100,000 population | 0.02 (0.13) | 0.0 | 0.0 | 0.017 | -0.668 – 0.703 | 0.96 |
| Community health centers* | Per 100,000 population | 9.4 (22.2) | 0.0 | 23.9 | 0.003 | -0.001 – 0.007 | 0.17 |
| Pharmacies | Per 100,000 population | 10.6 (11.3) | 0.0 | 24.2 | 0.041 | 0.033 – 0.49 | <0.01 |
| Medically underserved area (All or part of county) | Yes | 2,796 (87.4%) | No | Yes | 0.635 | 0.360 – 0.911 | <0.01 |
|  | No | 402 (12.6%) |  |  | ref |  |  |
| Primary Care Physician per population | Per 10,000 population | 5.7 (4.3) | 1.4 | 10.7 | -0.139 | -0.160 – -0.119 | <0.01 |
| Neurologist | Per 10,000 population | 0.2 (0.6) | 0 | 0.6 | -0.128 | -0.290 – 0.034 | 0.12 |
| Radiologist | Per 10,000 population | 0.1 (0.4) | 0 | 0.4 | -0.187 | -0.436 – 0.062 | 0.14 |
| Internists | Per 10,000 population | 1.5 (2.2) | 0 | 3.8 | -0.054 | -0.097 – -0.012 | 0.01 |
| Environmental features |  |  |  |  |  |  |  |
| Air quality | Annual PM2.5 level (µg/m³) | 7.5 (1.7) | 5.0 | 9.4 | 0.763 | 0.717 – 0.810 | <0.01 |
| Elevation above sea level | Mean elevation in 100 feet | 13.1 (14.9) | 1.0 | 33.9 | -0.088 | -0.093 – -0.083 | <0.01 |
| Access to parks** | Percent of population | 32.1 (27.2) | 2.1 | 74.1 | -0.020 | 0.023 – -0.016 | <0.01 |
| Mobility | Percent population that moved in the past year | 11.3 (3.9) | 7.0 | 16.0 | -0.002 | -0.021 – -0.025 | 0.87 |
| Drinking water violations | Yes | 1,204 (37.7%) | No | Yes | 0.027 | -0.216 – 0.162 | 0.78 |
|  | No | 1,994 (62.3%) |  |  | ref |  |  |
| Crowding | percent households with >1 person per room | 2.5 (2.2) | 0.7 | 4.7 | -0.009 | -0.049 – 0.032 | 0.684 |
| Food environmental index*** | County wide index | 7.6 (1.3) | 6.0 | 9.0 | -0.389 | -0.460 – -0.318 | <0.01 |
| Socioeconomic status |  |  |  |  |  |  |  |
| Median household income | In the year 2019 in 1000 US dollars | 62.9 (16.3) | 45 | 82 | -0.041 | -0.046 – -0.035 | <0.01 |
| Unemployment rate | Percent of population | 3.7 (1.4) | 2.3 | 5.2 | 0.356 | 0.290 – 0.422 | <0.01 |
| Medicare advantage penetration | Percent of >65 population in 2019 | 30.7 (16.8) | 8 | 50.13 | 0.025 | 0.019 – 0.030 | <0.01 |
| Uninsured rate | Percent of <65 population | 11.7 (5.5) | 6 | 19.5 | 0.022 | 0.005 – 0.039 | 0.01 |
| Living in poverty | Percent of population | 14.8 (5.8) | 8.6 | 22.4 | 0.149 | 0.134 – 0.164 | <0.01 |
| On supplemental nutritional assistance | Percent of population | 13.4 (7.4) | 5.2 | 23.8 | 0.121 | 0.109 – 0.132 | <0.01 |
| Broadband internet coverage | Percent of population | 82.1 (7.6) | 72.6 | 90.5 | -0.060 | -0.072 – -0.048 | <0.01 |
| Less than high school education | Percent of population | 11.9 (5.9) | 5.7 | 19.8 | 0.121 | 0.106 – 0.136 | <0.01 |
| Less than college education | Percent of population | 76.5 (10.0) | 63.1 | 86.5 | 0.072 | 0.063 – 0.081 | <0.01 |
| Primary economic sector type**** | Nonspecific | 1,255 (39.2%) | Nonspecific | Recreation | ref |  |  |
|  | Farming | 444 (13.9%) |  |  | -2.026 | -2.398 – -1.753 | <0.01 |
|  | Mining | 224 (7.0%) |  |  | -0.645 | -1.002 – -0.287 | <0.01 |
|  | Manufacturing | 505 (15.8%) |  |  | 0.091 | -0.169 – 0.351 | 0.49 |
|  | Government | 409 (12.8%) |  |  | -0.404 | -0.684 – -0.123 | <0.01 |
|  | Recreation | 361 (11.3%) |  |  | -1.838 | -2.132 – -1.543 | <0.01 |
| GDP/capita | In thousands of US dollar | 61.8 (104.1) | 24.7 | 89.1 | -0.004 | -0.005 – -0.003 | <0.01 |

^The unadjusted estimates were generated using a generalized linear mixed model with a random intercept on the county level.
^^All variable values are derived for the years of 2019 or 2020.
\*Community-based clinics that primarily focus on preventative and primary care services accessible to people of all ages regardless of insurance status.
\*\*Percentage of population living within half a mile of a park.
\*\*\*A healthy food environment measure from 0 (worst) to 10 (best) that accounts for both proximity to healthy foods and income.
\*\*\*\*Economic sector that employs the highest percentage of county residents.

Of the 61 included predictors, 51 (83.6%) were found to be significantly associated with stroke hospitalization rates (Table 1). The complete case analysis descriptive statistics were not different from the multiple imputed data (Table S2).

### Variation in Hospitalization Rates

The cohort of 3,198 counties and county-equivalents had a mean observed stroke hospitalization rate of 11.2 per 100,000 (SD= 2.6, IQR= 3.7, 10^th^ percentile= 7.7, 90^th^ percentile= 14.5) (Figure 1). The sequentially adjusted models yielded a mean predicted hospitalization rates that are similar to the observed rate with a slightly reduced variance. The between-county explained variation increased from a R^2^ of 0.35 in model 1 that adjusted only for demographics to a R^2^ of 0.57 in the fully adjusted model (Model 6).

**Figure 1:**
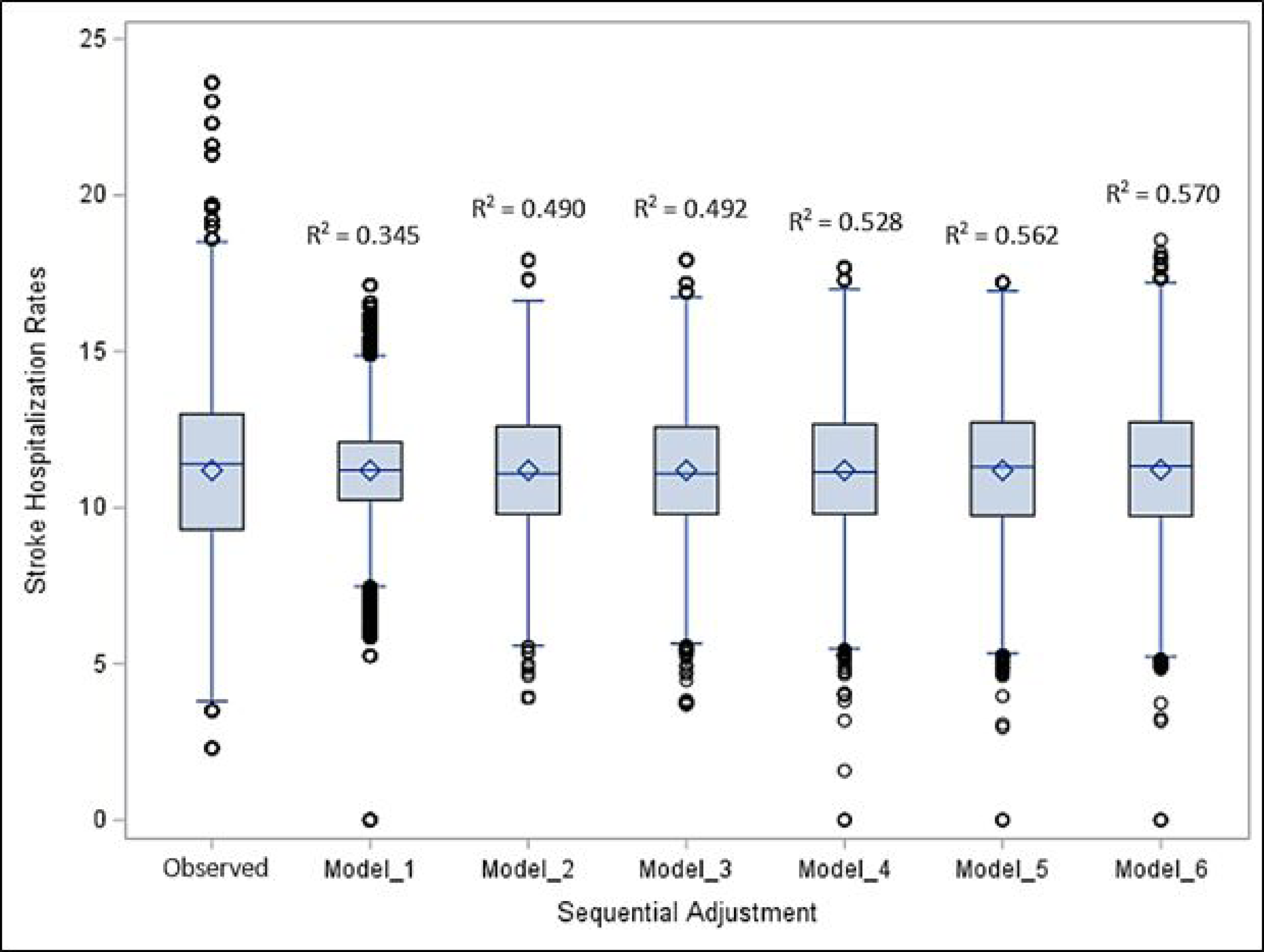
Box plot of sequentially adjusted models predicted stroke hospitalization rates per 100,000 population and their corresponding R^2^.* *Model 1= demographics (i.e., % Sex+ % age groups + % race groups + total population + rurality) Model 2= Model 1 covariates + population health panel/ stroke risk factors (i.e., % Hx CVD + % Hx high cholesterol + % Hx of high blood pressure + % Hx diabetes + % Hx of obesity + % physical inactivity + % smoking) Model 3= Model 2 covariates + treatment of risk factors (i.e., total Medicare reimbursement per enrollee + % nonadherence to blood pressure medication + % nonadherence to cholesterol lowering medication) Model 4= Model 3 covariates + healthcare delivery and access per population (i.e., # of hospitals + # of hospitals with neurological services + # of hospitals with CT + # of hospitals with tele stroke + # of certified stroke centers + # of hospitals + # of pharmacies + # of primary care physicians + # of radiologists + # of neurologists + # of internists + county includes medically underserved areas (yes/no)) Model 5= Model 4 covariates + environmental features (i.e., air quality + elevation above sea + access to park + mobility + drinking water violations + crowding + food index) Model 6= Model 5 covariates + socioeconomic status (i.e., household income + unemployment rate + Medicare advantage penetration + uninsured rate + % poverty + % internet coverage + % less than high school and college education + primary economic sector + GDP per population)

### Predictors of Stroke Hospitalization Rates

Our fully adjusted model had 17 out of 61 (27.9%) significantly associated predictors (Table 2 and Table S4). We found that the most impactful factors in predicting higher hospitalization rates in descending order included having a history of taking antihypertensive medication (a county at the 10^th^ percentile of antihypertensive medication prevalence had a predicted stroke hospitalization rate of 5.47 vs. 7.90 at the 90^th^ percentile [difference of 2.43]), smoking, non-adherence to antihypertensive medication, unemployment rate, total Medicare reimbursement per enrollee, air quality, number of internists per 10,000 population, percent on nutritional supplement assistance, and MA penetration rates (Table 2).

**Table 2:** The range of predicted hospitalization rates in the fully adjusted model using the 10th and 90th percentile value of the included predictors.*

| Category | Factor | Predicted change in expected hospitalization rate with covariate at the 10<sup>th</sup> percentile of the covariate level** | Predicted change in expected hospitalization rate with covariate at the 90<sup>th</sup> percentile of the covariate level** | Difference (90<sup>th</sup> -10<sup>th</sup>) | P-value |
| --- | --- | --- | --- | --- | --- |
| Demographics | Population aged 65-69 (%) | -0.63 | -1.09 | -0.46 | 0.04 |
|  | Rurality | 0.00 | -0.44 | -0.44 | <0.01 |
| Stroke risk factors/<br>population health panel | Coronary heart disease (%) | -3.27 | -5.54 | -2.28 | <0.01 |
|  | Hypertension (%) | 5.47 | 7.90 | 2.43 | <0.01 |
|  | Obesity (%) | -0.33 | -0.66 | -0.32 | 0.04 |
|  | Smoking (%) | 2.50 | 4.39 | 1.89 | <0.01 |
| Treatment of medical risk factors | Total Medicare reimbursement per enrollee | 0.99 | 1.42 | 0.43 | <0.01 |
|  | Medication nonadherence to antihypertensive medication (%) | 1.71 | 2.82 | 1.11 | <0.01 |
| Healthcare delivery and access | Number of hospitals per 100k population | 0.00 | -0.40 | -0.40 | <0.01 |
|  | Primary care physicians per 10k population | -0.08 | -0.60 | -0.53 | <0.01 |
|  | Internist per 10k population | 0.00 | 0.37 | 0.37 | <0.01 |
| Environmental features | Air quality (Annual PM2.5 level $\mu\text{g}/\text{m}^3$) | 0.46 | 0.87 | 0.41 | <0.01 |
|  | Elevation above sea level (in 100 feet) | -0.03 | -1.12 | -1.09 | <0.01 |
| Socioeconomic status | Unemployment rate (%) | 0.51 | 1.16 | 0.65 | <0.01 |
|  | Medicare Advantage penetration (%) | 0.05 | 0.29 | 0.25 | 0.04 |
|  | On nutritional supplemental assistance (%) | 0.12 | 0.55 | 0.43 | 0.02 |
|  | Less than college education (%) | -1.41 | -1.93 | -0.52 | <0.01 |

\*The most impactful variables were chosen from the pool of the significantly adjusted factors (n=17).
\*\* The 10<sup>th</sup> or 90<sup>th</sup> percentile predicted change in expected hospitalization rate = covariate 10<sup>th</sup> or 90<sup>th</sup> percentile\*model estimate of the covariate(example: 10<sup>th</sup> percentile prevalence coronary heart disease prevalence = 5.6, fully adjusted model covariate estimate = -0.583, predicted change = 5.6\*-0.584 = -3.27)

The most impactful factors in predicting lower stroke hospitalization rates in descending order included CHD (a county at the 10^th^ percentile of CHD prevalence had a predicted stroke hospitalization rate of −3.27 vs. −5.54 at the 90^th^ percentile [difference of −2.28]), the elevation of the county above sea level, number of primary care physicians per 10,000 population, proportion with less than college education, proportion ages 65-69 years old, rurality, number of hospitals per 100,00 population, and proportion with obesity (Table 2). Using the fully adjusted model, about 49.1% and 50.9% of the examined counties had above than expected and below than expected hospitalization rates, respectively. Counties in the northwest and New England (far northeast) region generally had lower-than-expected stroke hospitalization rates (Figure 2).

**Figure 2:**
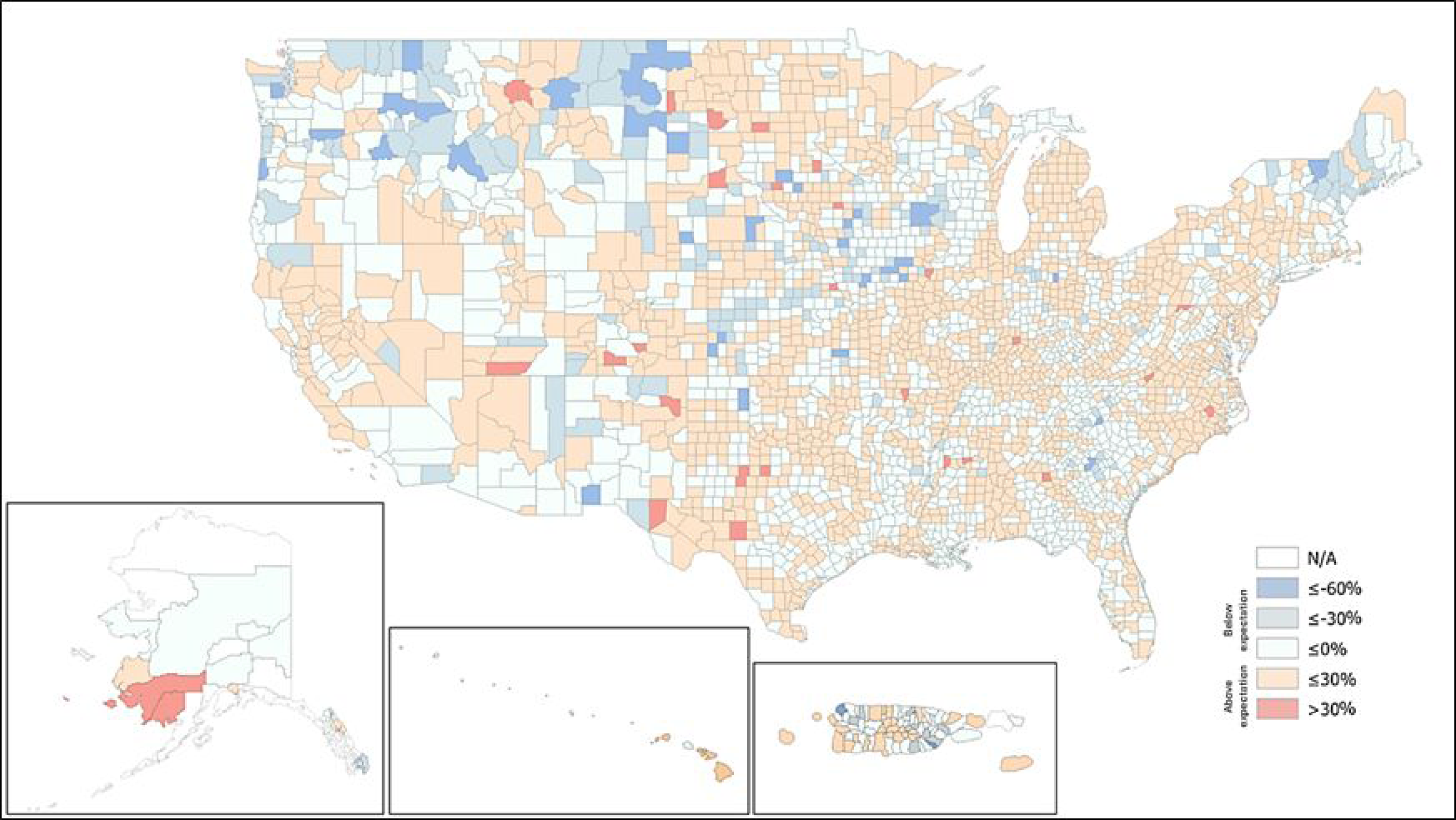
Proportional differences between crude and predicted hospitalization rates at the county level.* *Proportional difference= ((observed hospitalization rate – predicted hospitalization rate)/observed hospitalization rate)x100%

## Discussion

In this hypothesis-generating national cross-sectional study of county-level stroke hospitalization rates among Medicare FFS beneficiaries, we found that county level demographics, population stroke risk factor health panel, treatment, healthcare delivery and access, environment, and socioeconomic characteristics predicted a substantial proportion (R^2^= 0.57) of variation in county level hospitalization rates. Our findings support the consensus that place of residence is strongly associated with stroke hospitalization and identifies some potential stroke risk factors of interest as well as suggests some possible opportunities to improve stroke systems.^9^ These findings at the county-level may be able to inform public health programs and policies, as such policies are commonly designed and implemented at the local level.

We found that most social determinants of health examined were associated with stroke hospitalization rates in the expected direction. However, the patterns observed for rurality, MA penetration, education, race, and region of residence warrants further investigation. First, the inverse association between rurality and hospitalization may reflect reduced access to care, higher pre-hospital mortality (selective survival bias), or potentially protective environmental exposures such as lower air pollution or active lifestyle.^12^ Second, the positive association between MA penetration and hospitalization may reflect underlying selection effects as MA plans disproportionately enroll lower-income, Black or Hispanic individuals, those with poorer health status, and those with fewer years of education driven by lower or zero premiums and supplemental benefits.^13^ In addition, MA plans tend to code diagnosis more aggressively compared to FFS, reflecting coding intensity rather than true differences in morbidity.^14^ To our knowledge, no prior study has directly compared stroke hospitalization rates between MA and FFS beneficiaries; we hypothesize that the combined demographic, health, and socioeconomic profile of MA enrollees may contribute to increased hospitalization. Third, although higher educational attainment is generally associated with reduced stroke incidence,^15^ our analysis demonstrated the opposite. We hypothesize that this unexpected inverse association may be attributable to lower stroke literacy leading to delayed or missed diagnoses,^16^ to a differential access to care with the possibility of underdiagnosis amongst individuals with less education or overdiagnosis amongst those with more education.^17^ Fourth, despite well-documented geographic and racial disparities in stroke incidence—including the “stroke belt” and higher rates among Black compared with White individuals^18, 19^—none of the county-level demographic variables were associated with hospitalization after multivariable adjustment, suggesting that to the extent that regional demographics influence hospitalization, they are likely mediated through other factors captured in the model. Finally, our observation that the Northwest and New England regions exhibit lower-than-expected hospitalization rates is consistent with Schieb et al., who reported persistently lower hospitalization clusters in these areas based on FFS data from 1995–2006.^9^ Historically, these regions have the most favorable socioeconomic and healthcare profiles, yet no comprehensive analysis have identified the underlying protective factors.^9, 20, 21^

We also found, aligned with prior expectations, that conventional risk factors and risk factor treatment including hypertension, smoking, and nonadherence to antihypertensive therapy were independently associated with increased hospitalization rates. Both hypertension and smoking are established modifiable risk factors for stroke.^22^ Poor adherence to antihypertensive medication is also linked to a markedly higher short- and long-term stroke incidence, demonstrating a dose–response effect whereby lower adherence confers greater hospitalization risk.^23^ The negative association between obesity and hospitalization was not as clearly aligned with our expectations. While obesity is not considered an independent stroke risk factor as its effect are largely mediated by metabolic sequelae such as hypertension, diabetes, and hypercholesterolemia,^24^ it was not expected that regions with higher obesity rates would have lower stroke hospitalization rates. Our findings are novel and raise potential hypotheses: 1) receiving an obesity diagnosis may be a marker of health care access and 2) adherence to management of obesity-related metabolic disorders may mitigate stroke-related hospitalizations.

We also identified several unexpected and harder to explain findings related to history of CHD, total Medicare reimbursements for cardiovascular disease encounters, number of hospitals per population and average altitude above sea level. Notably, we observed an apparently contradictory association between history of CHD that was associated with lower hospitalization rates and total Medicare reimbursement for cardiovascular diseases that was associated with higher hospitalization rates. Several explanations are possible. In spite of the fact that CHD confers a twofold higher risk of stroke,^25^ because individuals with CHD tend to experience stroke at an older age than those without CHD,^22, 26^ CHD diagnosis and management prior to stroke may result in greater healthcare intensity, attenuating the observed hospitalization risk in this group.^27^ However, the possibility that more intense healthcare is associated with lower stroke hospitalization rates is contradicted by the positive association between total Medicare reimbursements and stroke hospitalization. The tension between those opposing explanations suggest a knowledge gap that warrants further study. In terms of the positive association with number of hospitals, scarce evidence suggest that an increase in the number of hospitals and providers may increase stroke hospitalizations by improving stroke diagnosis, decrease missed strokes or resulting in increased false positive stroke diagnosis.^9, 28^ Finally, the protective association with altitude conformed with findings from two reviews and meta-analyses encompassing studies from the U.S., Switzerland, Greece, China, and India.^29, 30^ Additionally, being born at higher versus lower altitude appears to exert an independent, additive protective effect.^31^ However, there is no clear understanding of the mechanisms leading to the altitude protective effect.

## Limitations

Studying hospitalization rates at the county level may be insensitive to important regional factors that exist at more granular geographic levels.^5^ This potential limitation may be partially offset by the idea that local policy levers of action (i.e., governmental or administrative regions) often exist at the county level. Further, our approach does not account for the patients with stroke who are not hospitalized and will underestimate trends in regions with more outpatient stroke diagnoses and overestimate in regions with a higher proportion of patients diagnoses. Although 75% of all stroke hospitalizations occur among people aged ≥65,^32^ by relying on Medicare data, our results are insensitive to regional differences in stroke in the young. Further, our hospitalization rates are underestimated because we included only FFS without MA data, this is likely partially offset by including MA penetration rates in the model.

Considerable but lower-than-expected unexplained county-level variance in stroke hospitalization rates exists after accounting for a wide variety of known and potential predictors. The findings of this study may inform future work to understand regional variation in stroke hospitalization rates and assist the development of targeted nuanced interventions to regions which have higher-than-expected stroke hospitalization rates and identify regions with lower-than-expected rates as a source of assessment to look for protective factors.

## Data Availability

Data is publicly available through the CDC’s Interactive Atlas of Heart Disease and Stroke (ATLAS)

http://nccd.cdc.gov/DHDSPAtlas

## Acknowledgments

None

